# ECHOCARDIOGRAPHY ABNORMALITIES IN PREECLAMPSIA WITH SEVERE FEATURES

**DOI:** 10.64898/2026.06.14.26355587

**Authors:** Ashakiran Thavarsingh Rathod, Sarojini, Ranga PC Sree, S Prathima, Sukeshini Deshmukh, Syeda Aliya Fathima

## Abstract

**Purpose:** To determine the frequency of echocardiographic abnormalities in women with preeclampsia with severe features.

To describe the spectrum and types of echocardiographic abnormalities associated with preeclampsia with severe features.

**Method:** This is a Prospective observational study conducted in Vani Vilas hospital attached to Bangalore Medical College and Research Institute, Bangalore from January 2023 to December 2025. 560 pregnant women diagnosed with severe preeclampsia(SPE) were included in the study. Chronic hypertension without superimposed preeclampsia, underlying cardiac diseases and previous history of peripartum cardiomyopathy were excluded from the study. Transthoracic echocardiography-TTE (2D ECHO) was done to evaluate cardiac structure and function. Echocardiographic abnormalities identified during the study were documented and analysed using descriptive statistical methods.

**Results:** Abnormalities in ECHO was noted in 23.03%. A unique finding was the documentation of elevated pulmonary artery systolic pressures (PASP) suggestive of Pulmonary Hypertension (PH) (PASP >35 mm HG) among 20.25% of the participants. It was also the commonest abnormality on ECHO. Mild PH was the commonest (15.71%), moderate PH was seen in 3.92% and severe PH in 0.71% of cases. Next most frequent abnormality was moderate to severe valvular regurgitation (10%), followed by left ventricular hypertrophy (5.53%). Diastolic dysfunction (DD) was seen in 3.92%, systolic dysfunction(SD) in 3.57%, chamber dilatation in 3.57% and LV global hypokinesia in 3.03% cases of SPE

**Conclusion:** Preeclampsia with severe features (SPE) is associated with 23.03% abnormalities on echocardiography. SPE is associated with systolic dysfunction, diastolic dysfunction, chamber dilatation, valvular regurgitation, left ventricular hypertrophy and pulmonary hypertension.

## Introduction

Preeclampsia is a multisystem disorder that significantly affects the cardiovascular system. The cardiac involvement in preeclampsia is complex and results from hemodynamic alterations, endothelial dysfunction, hypertension and myocardial remodelling^1^.

In preeclampsia, there is increased afterload due to hypertension. The expected physiological blood volume expansion does not happen leading to intravascular volume depletion when compared to normal pregnant women^2^. Administration of intravenous crystalloids in such patients can therefore abruptly increase preload and precipitate cardiac decompensation.

Endothelial dysfunction, a hallmark of preeclampsia, contributes leakage of intravascular fluid into the extracellular compartment, and hemoconcentration. Additionally, reduced serum albumin levels lower colloid oncotic pressure, further promoting pulmonary and systemic fluid extravasation^1^.

Cardiac output in preeclampsia is often reduced secondary to increased peripheral vascular resistance^3^. Serial echocardiographic studies have demonstrated evidence of diastolic dysfunction in women with preeclampsia^4,5^. This dysfunction may be particularly detrimental in patients with pre-existing left ventricular hypertrophy due to chronic hypertension.

Peripartum cardiomyopathy, has a recognized association with preeclampsia, with approximately 22% of cases coexisting with preeclampsia^6^.

Elevated levels of high-sensitivity cardiac troponin and NT-proBNP have also been reported in women with preeclampsia^7,8^.

Acute pulmonary edema complicates nearly 5.6% of preeclampsia cases and remains an important cause of maternal morbidity and mortality^9^. It may result from one or a combination of increased afterload, endothelial dysfunction, capillary leak, diastolic dysfunction, and altered preload conditions.

Recent studies have also demonstrated worsening abnormalities in global longitudinal strain (GLS) among women with preeclampsia. These changes also correlate with circulating antiangiogenic biomarkers such as soluble fms-like tyrosine kinase-1 (sFlt-1) and soluble endoglin (sEng) further supporting the concept of myocardial involvement in preeclampsia^10^.

Echocardiographic evaluation in severe preeclampsia can identify structural and functional cardiac abnormalities and provide clinically valuable information regarding fluid therapy, anaesthetic management, antihypertensive selection, and anticoagulation strategies. Early identification of abnormal cardiac remodelling may also facilitate initiation of long-term cardioprotective therapy in the postpartum period.

Despite these recognized cardiovascular risks, routine screening with transthoracic echocardiography has not yet been universally standardized in the management of severe preeclampsia. Most available studies evaluating echocardiographic abnormalities in preeclampsia are limited by small sample size. The present study was designed to address this gap in knowledge and to characterize the frequency and spectrum of echocardiographic abnormalities in 560 women with severe preeclampsia at a single tertiary care centre.

### Objective

To determine the frequency of echocardiographic abnormalities in women with preeclampsia with severe features.

## Method

This is a Prospective observational study conducted in Vani Vilas hospital attached to Bangalore Medical College and Research Institute, Bangalore from January 2023 to December 2025. Vani Vilas hospital is a tertiary care referral hospital. A total of 560 pregnant women diagnosed with severe preeclampsia were included in the study.

The criteria used for the diagnosis of severe preeclampsia (SPE) were as follows.

1. Blood pressure more than or equal to 160/110 mm Hg Or Hypertension (more than or equal to 140/90 mm Hg) with one of the following
2. Platelet count less than 100,000/µL
3. Elevated liver enzymes
4. Renal failure
5. Acute pulmonary edema
6. Symptoms such as headache, blurring of vision, convulsions, epigastric burning and reduced urine output
7. Fetal growth restriction

Chronic hypertension without superimposed preeclampsia, underlying cardiac diseases and previous history of peripartum cardiomyopathy were excluded from the study. Written informed consent was obtained from all women participating in the study. Detailed history taking, clinical examination, and relevant investigations were performed in all cases. Transthoracic echocardiography-TTE (2D ECHO) was conducted to evaluate cardiac structure and function. The clinical and echocardiographic findings were recorded in a standardized proforma. Echocardiographic abnormalities identified during the study were documented and analysed using descriptive statistical methods.

## Results

The study included 560 pregnant women diagnosed with preeclampsia with severe features.

The predominant age group was 18–25 years (52.5%), followed by 26–30 years (26.42%)(table 1).

**Table 1:** Age distribution.

| Age | Number N=560 | Percentage |
| --- | --- | --- |
| <18 | 01 | 0.17 |
| 18-25 | 294 | 52.5 |
| 26-30 | 148 | 26.42 |
| 31-35 | 83 | 14.82 |
| 36-40 | 30 | 5.35 |
| 41-45 | 04 | 0.71 |

Regarding parity, primigravida women constituted the largest proportion of the cohort (46.60%), while those in their second pregnancy accounted for 27.5%(table 2).

**Table 2:** Parity distribution.

| Parity | Number N=560 | Percentage |
| --- | --- | --- |
| 1<sup>st</sup> pregnancy | 261 | 46.60 |
| 2<sup>nd</sup> | 154 | 27.5 |
| 3<sup>rd</sup> | 112 | 20 |
| 4<sup>th</sup> | 23 | 4.10 |
| 5<sup>th</sup> | 06 | 1.07 |
| 6<sup>th</sup> | 02 | 0.35 |
| 7<sup>th</sup> | 02 | 0.35 |

Most of the participants (66.85%) presented at preterm gestational ages, while 33.15% were at term. The most frequent gestational age range being 34–37 weeks (25.75%), followed by 37–39 weeks (22.85%)(table 3).

**Table 3:** Gestational age.

| Gestational age in weeks | Number N=560 | Percentage |
| --- | --- | --- |
| Less than 24 | 05 | 0.89 |
| 24 to 28 | 35 | 6.25 |
| 28 to 32 | 103 | 18.39 |
| 32 to 34 | 87 | 15.53 |
| 34 to 37 | 144 | 25.75 |
| 37 to 39 | 128 | 22.85 |
| 39 to 42 | 56 | 10 |
| more than 42 | 02 | 0.35 |

The distribution of blood pressure (BP) at the time of admission revealed a significant range of hypertensive severity within the cohort. Notably, 32.85% (n=184) of the women presented with BP in the 160/110–179/119 mmHg range, representing the largest single group in the study(table 4).

**Table 4:** BP on admission.

| BP on Admission | Number N=560 | Percentage |
| --- | --- | --- |
| <140/90 | 114 | 20.35 |
| 140/90 - 149 by 99 | 117 | 20.89 |
| 150/100 – 159/109 | 96 | 17.14 |
| 160/110 -179/119 | 184 | 32.85 |
| 180/120-199/139 | 36 | 6.42 |
| 200/140-239/159 | 12 | 2.10 |
| >240/160 | 01 | 0.17 |

Extreme hypertension was also documented: 6.42% (n=36) of patients presented with BP between 180/120 and 199/139 mmHg, while 2.10% (n=12) were in the 200/140–239/159 mmHg category. One patient (0.17%) presented with a critical BP exceeding 240/160 mmHg. Collectively, nearly 41.5% of the participants exhibited blood pressure levels ≥160/110 mmHg on admission. Moderate high BP readings in the range of 150/100 and 159/109 mmHg were seen in 17.14% (n=96).

20.35% (n=114) of the patients presented with BP levels below 140/90 mmHg at the time of admission. This relatively lower recording in a cohort diagnosed with preeclampsia with severe features is likely attributable to the administration of antihypertensive medications at primary or secondary care facilities prior to their referral to our tertiary institute.

The HDP spectrum at presentation was most commonly preeclampsia with severe features (76,60%). 14.28% presented with eclampsia. 9.10% presented with preeclampsia with severe features superimposed on chronic hypertension(table 5).

**Table 5:**
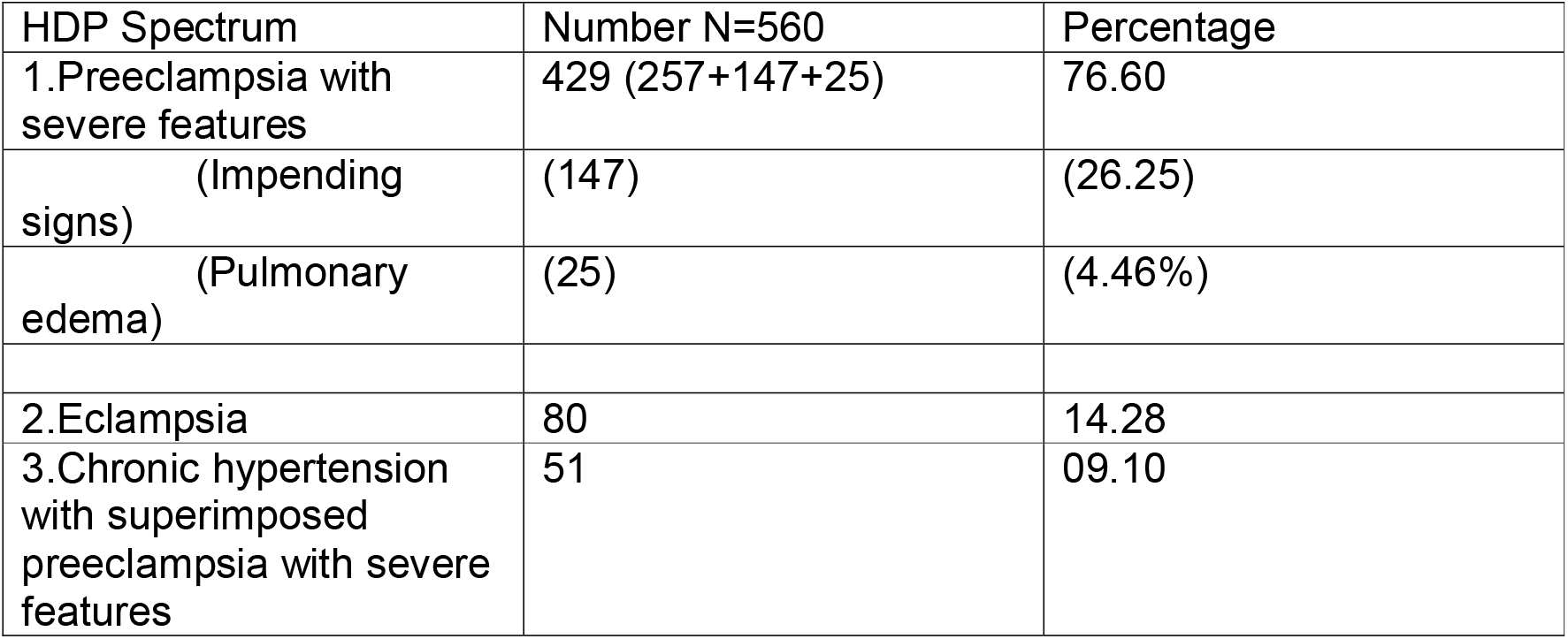
HDP spectrum at presentation.

| HDP Spectrum | Number N=560 | Percentage |
| --- | --- | --- |
| 1.Preeclampsia with severe features | 429 (257+147+25) | 76.60 |
| (Impending signs) | (147) | (26.25) |
| (Pulmonary edema) | (25) | (4.46%) |
| 2.Eclampsia | 80 | 14.28 |
| 3.Chronic hypertension with superimposed preeclampsia with severe features | 51 | 09.10 |

Out of 560 women, 23.03%(N=129) had abnormal ECHO. 76.96% (N=431) had normal ECHO. Among the normal are also included women showing mild abnormalities like mild valvular regurgitation in 27.50% (N=154).

87 (15.53%) women were admitted to the Obstetric intensive care unit (ICU) for various indications. Among them 37(42.52%) had normal ECHO and 50(57.47%) had abnormal ECHO.

Abnormalities on Echocardiography(table 6).

**Table 6:** Echocardigraphy abnormalities.

|  | N=560 | Percentage |  |  |
| --- | --- | --- | --- | --- |
| Abnormal | 129 | 23.03% |  |  |
| Mild abnormalities(mild valvular regurgitation included as normal ECHO) | 154 | 27.50% |  |  |
| Abnormal ECHO parameter | Number among total PE with severe features N=560(%) | Percentage among SPE | Abnormal ECHO N=129 | Percentage among abnormal ECHO |
| **Systolic dysfunction** | 20(based on Ef<55) | 3.57 | 20 | 15.50 |
| Chamber dilatation | 20 (3.57) | 3.57 | 20 | 15.50 |
| LA (left atrium) | 17 | 3.03 | 17 | 13.17 |
| LV (left ventricle) | 14 | 2.50 | 14 | 10.85 |
| RA (right atrium) | 11 | 1.96 | 11 | 08.52 |
| RV (right ventricle) | 08 | 1.42 | 08 | 06.20 |
| LV global hypokinesia | 17 | 3.03 | 17 | 13.17 |
| Ejection fraction < /=45 | 14 | 2.50 | 14 | 10.85 |
| Ejection fraction<br>46 to 55 | 06 | 1.07 | 06 | 04.65 |
| **Diastolic**<br>**dysfunction** | 22 | 3.92 | 22 | 17.05 |
| Grade 1 | 11 | 1.96 | 11 | 08.52 |
| Grade 2 | 10 | 1.78 | 10 | 07.75 |
| Grade 3 | 01 | 0.17 | 01 | 00.77 |
| **Left ventricular**<br>**hypertrophy** | 31 | 5.53 | 31 | 24.03 |
| **Pulmonary**<br>**hypertension**<br>**Elevated -**<br>**Pulmonary**<br>**artery systolic**<br>**pressure (PASP**<br>**mm Hg)** | 114 | 20.35 | 114 | 88.37 |
| Mild PH<br>(PASP>35 -50) | 88 | 15.71 | 88 | 68.21 |
| Moderate PH<br>(PASP>50-70) | 22 | 3.92 | 22 | 17.05 |
| Severe PH<br>(PASP>=70) | 04 | 0.71 | 04 | 03.10 |
| Valvular<br>regurgitation | 445<br>(56 mod to<br>Sev)<br>(389 mild) | 79.56%<br>(10%)<br>(69.46%) | -<br>56<br>- | -<br>43.49<br>- |
| Moderate to<br>severe mitral<br>regurgitation | 24 | 4.28% | 24 | 18.60 |
| Moderate to<br>severe tricuspid<br>regurgitation | 32 | 5.71 | 32 | 24.80 |
| Mild mitral<br>regurgitation | 186 | 33.21 | - | - |
| Mild tricuspid<br>regurgitation | 190 | 33.92 | - | - |
| Mild aortic<br>regurgitation | 13 | 2.32 | - | - |

A unique finding in this study was the documentation of elevated pulmonary artery systolic pressures (PASP) suggestive of Pulmonary Hypertension (PH) (PASP >35 mm HG) among 20.25% of the participants. More striking is that it was also the commonest abnormality on ECHO. Mild PH was the commonest (15.71%), moderate PH was seen in 3.92% and severe PH in 0.71% of cases. Next most frequent abnormality was moderate to severe valvular regurgitation (10%), followed by left ventricular hypertrophy (5.53%). Diastolic dysfunction(DD) was seen in 3.92%, systolic dysfunction(SD) in 3.57%, chamber dilatation in 3.57% and LV global hypokinesia in 3.03% cases of preeclampsia.

## Discussion

It is “to our knowledge, one of the largest single-centre cohorts evaluating echocardiographic abnormalities in severe preeclampsia.” we observed diastolic dysfunction, systolic dysfunction, cardiac chamber dilatation, global hypokinesia, mitral and tricuspid valvular regurgitation, left ventricular hypertrophy and pulmonary hypertension. The large sample size of the present study enabled us to reliably estimate the frequency of these cardiac abnormalities in women with severe preeclampsia.

### Systolic dysfunction

In the present study, left ventricular systolic dysfunction(LVSD) was defined as a left ventricular ejection fraction (LVEF) of less than 55%. Using this criterion, SD was identified in 3.57% of women with SPE. 2D TTE evidence of SD may include a reduced ejection fraction, global hypokinesia, regional wall motion abnormalities, and reduced fractional shortening^11^.

Cardiac dysfunction in preeclampsia has traditionally been described predominantly by abnormalities of diastolic function^12,13,14,15^. However, emerging evidence suggests that systolic impairment may also occur in this population. Several studies have demonstrated evidence of impaired systolic function in women with preeclampsia^10.16^. Our findings add to the growing body of evidence supporting the presence of systolic dysfunction in preeclampsia.

In the present study with conventional echocardiography 3.57% were reported to have reduced LVEF. This may be just the tip of an iceberg. There could be cases with SD with normal EF (Subclinical systolic dysfunction). This subclinical dysfunction may be picked up by 3D speckle tracking echocardiography as demonstrated by Shahul S et al^10^.

Global hypokinesia was seen in 3.03% cases which suggest systolic dysfunction. Chamber dilatation was seen in 3.57% of cases which suggests cardiac muscle remodelling. Left ventricle and left atria were dilated in 2.50% and 3.03% of all cases respectively. Literature usually has studies evaluating the left heart. Few studies show involvement of the right heart also^17^, which has been observed in the present study where right atria and right ventricle were dilated in 1.96% and 1.42% of cases respectively indicating a right ventricular remodelling.

A cross-sectional study was done in Chennai, India involving 80 pregnant women (40 hypertensive and 40 normotensive). Echocardiography was done in all 80 women. Results revealed significant left ventricular chamber dilatation with wall thickening(remodelling), impaired diastolic relaxation patterns and left atrial enlargement. These are very similar to our findings. However, they found the systolic function was preserved^18^. This may be due to the small sample size.

Cong J and colleagues studied 43 women with early onset and 41 women with late onset preeclampsia by 3D speckle-tracking echocardiography and compared with 81 normotensive pregnant women. They reported significant eccentric hypertrophy, ventricular systolic and diastolic dysfunction, left atrial remodelling and left atrial function abnormalities^16^. Left atrial remodelling is more frequently described in the literature which also correlates with our study where we report left atrial dilatation as the most common among chamber dilatation.

#### Valvular regurgitation

Moderate to severe valvular regurgitation was noted in 10% cases. Tricuspid regurgitation was seen in 5.71% and Mitral regurgitation in 4.28% of SPE. We also noted mild valvular regurgitation very frequently which accounted to 69.49% cases, but we have not taken them into account as an abnormal finding. Valvular regurgitation occurs secondary to the chamber dilatation which again suggests SD and cardiac remodelling.

### Diastolic dysfunction and left ventricular hypertrophy

We observed left ventricular diastolic dysfunction in 3.92% percent of cases. Most common was grade 1 DD followed by grade 2 and 3. The frequency of diastolic and systolic dysfunction (3.92% and 3.57%) was almost similar, with DD occurring only slightly more frequently than SD. We observed left ventricular hypertrophy in 5.53% of cases.

The prevalence of diastolic dysfunction reported in previous studies ranges from 12.7% to 40%, with reported frequencies of 12.7% by Vaught AJ et al^5^., 20% by Muthyala T et al^19^., and 40% by Melchiorre K et al^14^. These estimates are higher than the prevalence observed in the present study,

A comparative study among three groups was done by Dennis A T and colleagues, 40 preeclampsia, 40 healthy pregnant and 20 non-pregnant women. The hemodyanamics was studied using TTE. They concluded that preeclampsia is associated with increased cardiac output, vasoconstriction, increased ionotrophy and reduced diastolic function^12^.

Castleman JS et al in the systematic review of 36 studies involving 745 gestational hypertension and 815 PE concluded that both are associated with increased left ventricular mass. DD and LV remodelling was marked in early onset and severe PE. These findings were also associated with adverse outcomes^13^.

Melchiorre K and colleagues reported that echocardiographic abnormalities in preeclampsia have important implications for peripartum hemodynamic and intravascular volume management. The authors observed that women with global diastolic dysfunction are at an increased risk of developing acute pulmonary edema^15^.

Vaught AJ et al in a prospective observational study of 63 PEC (preeclampsia with severe features) women and 36 pregnant control women observed higher right Ventricular Systolic Pressure(RVSP) and decreased Right Ventricular Longitudinal Systolic Strain (RVLSS) for the right heart parameters. For left heart parameters, there was a difference in left atrial area size and in the thickness of posterior and septal wall. These findings indicate that preeclampsia is associated with structural and functional cardiac alterations involving both the right and left sides of the heart as also observed in the present study. They reported pulmonary edema in 9.5% of PEC^15^. In our study the frequency of pulmonary edema was 4.46%.

Rafik Hamad R and colleagues compared the echocardiography findings and doppler tissue imaging of the heart among 35 PE and 30 normal pregnant women. They reported left ventricular diastolic dysfunction in early onset preeclampsia and increased NT pro BNP and cystatin C when compared to normal pregnant women^20^.

### Long term effects

Women with preeclampsia were subjected to echocardiography 12 to 18 months after delivery. They were then followed in their next pregnancy. 29% of women developed recurrent preeclampsia. These women with recurrent preeclampsia had signs of diastolic dysfunction and increased left ventricular mass index when the ECHO was done in the non-pregnant state. This can be used as a prediction for recurrent preeclampsia^21^.

Vaught AJ et al studied 33 women with preeclampsia with severe features. Follow up was done for 4 years. At 4 years 48% developed hypertension within 4 years of delivery. These women had thicker left ventricular posterior walls on the antenatal Echocardiogram at the time of PEC diagnosis^22^. This suggests that women who have abnormal echo should be followed up aggressively for development of hypertension.

Although long-term follow-up data were not available in the present study, the identification of left ventricular diastolic dysfunction and ventricular hypertrophy in present study suggests the need for long-term cardiovascular surveillance.

### Pulmonary hypertension (PH)

Classified into 5 types based on the pathophysiology, hemodynamics, clinical features and management by WHO.

1. Pulmonary artery hypertension (PAH) 2. PH due to left heart disease 3. PH due to lung disease/hypoxia 4. Chronic thromboembolic PH (CTEPH) 5. Miscellaneous/multifactorial^23^.

In the present study, elevated pulmonary artery systolic pressure (PASP >35 mmHg) was observed in 114 of 560 women (20.35%) suggestive of pulmonary hypertension making it the most common echocardiographic abnormality identified in the cohort. Mild PH was present in 88 women (15.71%), while moderate and severe PH were observed in 22 (3.92%) and 4 (0.71%) women, respectively.

The probable explanation of elevated PASP seen in SPE could be as follows

1. Secondary to left heart changes
2. Secondary to the systemic inflammation associated with preeclampsia
3. Remodelling of the pulmonary arteries
4. Vasospasm of pulmonary arteries
5. Secondary to reduced GFR and renal dysfunction seen in SPE and metabolic products (toxin) accumulation

To our knowledge, pulmonary hypertension has not been widely reported as a predominant echocardiographic finding in women with severe preeclampsia.

While the association between preeclampsia and systemic hypertension is well established, the involvement of the pulmonary circulation in women with severe preeclampsia has received relatively limited attention. This observation may represent an underrecognized manifestation of severe disease. The presence of PH places these women at higher risk for right-sided heart failure and maternal collapse

A review of the available literature identified studies that have reported elevated pulmonary artery pressures on echocardiography in women with preeclampsia^18,24^. However, we were unable to identify studies in which pulmonary hypertension was described as a common echocardiographic finding or a distinct cardiovascular manifestation in a large cohort of women with severe preeclampsia.

## Conclusion

Preeclampsia with severe features(SPE) is associated with 23.03% abnormalities on echocardiography. SPE is associated with systolic dysfunction, diastolic dysfunction, chamber dilatation, valvular regurgitation, left ventricular hypertrophy and pulmonary hypertension.

### Strengths of the study

- Largest number from a single centre with respect to the sample size
- The study describes the spectrum of echocardiography abnormalities in severe preeclampsia and gives the frequency of each abnormality
- Includes preeclampsia with severe features
- Single centre which ensured homogeneity of the population
- Single centre which ensures echocardiography done by personnel with similar training.

### Limitation of the study

- Lack of a comparison group
- Lack of postpartum follow-up echocardiography, preventing assessment of whether the abnormalities were transient or persistent.
- Single-centre design, which is a strength for consistency but may limit generalizability to other populations

### Suggestions/recommendations

Routine ECHO in severe preeclampsia - Given the substantial burden of cardiovascular abnormalities identified in women with severe preeclampsia, echocardiographic evaluation should be considered as part of the comprehensive assessment of these patients, facilitating risk stratification, individualized clinical management, and appropriate postpartum cardiovascular follow-up. Further studies are warranted to evaluate the cost-effectiveness and clinical impact of routine echocardiographic screening in women with severe preeclampsia.

Marker of severity - The identification of pulmonary hypertension, systolic dysfunction, or diastolic dysfunction on echocardiography may represent evidence of advanced cardiovascular involvement in preeclampsia and could potentially serve as markers of severe disease. Further prospective studies are needed to validate their prognostic significance and evaluate their role in defining disease severity.

## Data Availability

all data produced in the present work are contained in the manuscript

## Acknowledgment

None

## Abbreviations

SPE: Preeclampsia with severe features is used interchangeably with severe preeclampsia in this article
PE: Preeclampsia
TTE: Transthoracic echocardiography
ECHO: Echocardiography
SD: Systolic dysfunction
DD: Diastolic dysfunction
LVEF: Left ventricular Ejection fraction
(PASP): Pulmonary artery systolic pressure
PH: Pulmonary hypertension

## Notes

### Competing Interest Statement

The authors have declared no competing interest.

### Author Declarations

Ethics Committee/IRB of Bangalore Medical College and Research Institute gave ethical approval for this work.

